# Household molecular epidemiology of *Streptococcus pyogenes* carriage and infection in The Gambia

**DOI:** 10.1101/2025.03.28.25324710

**Authors:** Gabrielle de Crombrugghe, Edwin P. Armitage, Alexander J. Keeley, Elina Senghore, Fatoumata Camara, Musukoi Jammeh, Amat Bittaye, Haddy Ceesay, Isatou Ceesay, Bunja Samateh, Muhammed Manneh, Gwenaëlle Botquin, Dalila Lakhloufi, Valerie Delforge, Saikou Y Bah, Jennifer N. Hall, Lionel Schiavolin, Claire E. Turner, Michael Marks, Thushan I. de Silva, Anne Botteaux, Pierre R. Smeesters, the MRCG StrepA Study Group

## Abstract

**Background:** High burden of *Streptococcus pyogenes (S pyogenes)* disease is seen in Africa which is also the continent with the least epidemiological data on circulating strains. We aimed to better characterise *emm*-types and *emm*-clusters associated with carriage and disease in a setting with high rheumatic heart disease (RHD) burden, a peri-urban area in The Gambia.

**Methods:** A one-year household cohort study was conducted in 2021-2022, recruiting 442 healthy participants from 44 households, looking for *S pyogenes* carriage and non-invasive infection. Pharyngeal and normal skin swabs were collected to assess carriage, pharyngitis and pyoderma swabs were captured to assess infection. Cultured isolates underwent *emm*-typing and were compared with a similar collection from 2018. Simpson’s reciprocal index (SRI) was used to measure diversity.

**Results:** 221 isolates showed a positive culture for *S pyogenes*, representing 52 *emm*-types and 16 *emm*-clusters, with 4 over-represented clusters comprising 65.2% of the isolates. *emm*-type diversity was high (SRI 29.3, 95% CI: 24.8-36.0). Looking at *emm*-type and intra-individual transmission, we found frequent transmission between pyoderma and intact skin, and evidence of bidirectional transmission between skin and pharynx in the same host. A comparison with pyoderma isolates collected in 2018 from the same region revealed no major changes in circulating *emm*-clusters.

**Conclusion:** This study provides the first molecular analysis of skin and throat isolates prospectively collected from carriage and non-invasive infection in Africa. In this RHD-endemic setting, pyoderma and skin carriage represent an important *S pyogenes* reservoir and should be included in further surveillance studies and public health interventions.

**Funding:** Wellcome Trust, FNRS (Belgium), ESPID

**Keypoints summary:**

- First molecular analysis of *S pyogenes* skin carriage in Africa. Skin carriage may be an important reservoir in RHD-endemic settings.
- Highest strain diversity is seen in pyoderma and lowest in pharyngitis.
- Four *emm*-clusters are dominant and stable over time.

## Background

*Streptococcus pyogenes* is among the top 10 leading causes of global infection-related morbidity and mortality worldwide and a WHO global priority for vaccine research and development (1, 2). Moreover, the recent 2022-2023 surge in invasive *S pyogenes* disease has attracted widespread attention and concern (3, 4). *S pyogenes* is responsible for a broad spectrum of clinical manifestations including asymptomatic carriage, superficial infections, invasive infections and immune-mediated sequelae such as acute rheumatic fever (ARF) and rheumatic heart disease (RHD) (5). ARF and RHD are responsible of 275 000 deaths each year in low-and middle-income countries (LMIC), the highest burden of disease being in sub-Saharan Africa (6, 7). However, epidemiological data from Africa are currently limited. A systematic review of global *S pyogenes* strain diversity identified 13 studies coming from Africa between 1990 and 2023, covering 8 countries and contributing only 3% of global isolates worldwide (table S1), and lacking both carriage and longitudinal epidemiological data (8). Robust regional and country-level data from the African continent are urgently needed to inform vaccine development. Epidemiologic surveillance of *S pyogenes* is based on whole genome sequencing and on the sequence of the *emm* gene coding for the surface M protein, called *emm*-typing (9). Currently, more than 270 *emm-types* have been described worldwide, which can be grouped into 48 *emm-clusters*, a functional classification based on both genetic relatedness and common biological properties (10). The diversity of circulating *emm*-types is known to be higher in LMIC compared to HIC (high income countries), and an inverse correlation is observed between strain diversity and the UNDP Human Development Index (8). There is increasing evidence that skin infection and asymptomatic throat carriage are important both as source of transmission and in pathological immune priming, leading to ARF and RHD (11–17).

The World Health Assembly has recognised that vaccine development against *S pyogenes* is a major public health priority (18). Current vaccine candidates can be broadly divided into M protein-based and non-M protein-based candidates, but only M protein-based candidates have currently reached human clinical trials yet (19). Based on available epidemiological data showing limited *emm*-type diversity in the USA and Europe, a 30-valent vaccine candidate targeting the M protein has been developed and shown to be immunogenic in humans (20, 21). However, concerns have been raised about its coverage in LMIC where the diversity of circulating *emm*-types is greater (22, 23). The aim of our study is to characterise the molecular epidemiology of *S pyogenes* carriage and infection isolates from a longitudinal household study conducted in a setting with high RHD burden, The Gambia (24), to assess epidemiological differences with a comparable cohort from 2018, and to evaluate the theoretical coverage of the 30 valent M-protein vaccine candidate (20, 21).

## Methods

### Study population and microbiology

Clinical isolates were obtained from a prospective study designed to determine the epidemiology of *S pyogenes* carriage and infection, natural immunity and factors influencing transmission dynamics within households (25). The study protocol has been published previously (26). This household cohort study was conducted in the peri-urban area of Sukuta, The Gambia, from August 2021 to September 2022. Briefly, a total of 44 households were recruited, resulting in 442 participants, aged 0-85 years. The cohort comprised 58% of children <18 years, with a median age of 15 years, 53% were female and the median household size was 7 people. Households underwent a baseline medical visit followed by 12 scheduled monthly visits. At monthly visits, participants provided an oropharyngeal swab and a composite normal skin swab from healthy skin surfaces on the arms, legs, and forehead to monitor asymptomatic carriage. Episodes of pharyngitis and pyoderma were also recorded at monthly visits, and wound swabs taken if pyoderma present. Participants presenting with a sore throat or skin lesions between scheduled monthly visits were seen at unscheduled visits and an oropharyngeal or wound swab was taken accordingly. A subgroup of participants underwent more frequent swabbing (supplementary material page 2 for details). Case definitions for pharyngitis, pyoderma, pharyngeal carriage and skin carriage were previously defined (25).

Swabs were placed in liquid Amies transport medium and kept in a cold box until culture the same day at Medical Research Council The Gambia (MRCG) microbiology laboratory. Swabs were plated on Colombia blood agar and beta-haemolytic colonies underwent latex agglutination testing (Prolex, Pro-Lab, Bromborough, UK) for Group A *Streptococcus*. Isolates positive for Group A *Streptococcus* were assumed to be *S pyogenes*. Isolates were stored at −80°C in 17% glycerol and shipped to the Molecular Bacteriology Laboratory in Brussels (ULB), Belgium, for *emm*-typing. Isolates were reconfirmed as Group A *Streptococcus* using latex agglutination (Pastorex, Bio-Rad, Marnes-la-Coquette, France), and underwent PCR-based *emm*-typing using the updated protocol (27). Sequences were analysed with the Centers for Disease Control (CDC) Streptococci Group A Subtyping Request form Blast 2.0 server (28). *Emm*-clusters were deduced from *emm*-typing results as previously described (10).

### Population-based analyses and intra-individual transmission

We included a single isolate per *emm*-type per month (31-day) and per individual to avoid inclusion of multiple isolates coming from the same infectious/carriage episode. Multiple isolates belonging to the same *emm*-type collected from the same individual within a 31-day interval were therefore excluded from population-based analyses (*emm*-type diversity, theoretical vaccine coverage and temporal comparison). However, all these “multiple isolates” were the focus of a specific analyses assessing intra-individual transmission, i.e. *emm*-type transmission between different niches in the same host. *S pyogenes emm*-type diversity was assessed by Simpson’s reciprocal diversity index (29).

### M-protein theoretical vaccine coverage

The 30-valent vaccine theoretical coverage was estimated using the published cross-opsonisation data, based on a killing level >50% by functional bactericidal assays (20, 21). Based on recent animal data on a 32-valent M protein vaccine candidate, we could estimate the direct coverage of this candidate but not the potential cross-opsonisation due to the lack of a clear cutoff regarding bactericidal activity (30).

### Temporal comparison of emm-types causing pyoderma

Pyoderma samples collected in 2021-2022 were compared with pyoderma samples collected in 2018 from the same area (Sukuta, The Gambia) (31–33). The population studied wasn’t the same over the two periods. The 2018 cohort included 1441 children under 5 years who were examined once between May and September 2018, and the 2021-2022 cohort included 442 participants (0-85 years) belonging to 40 households who were examined multiple times between August 2021 and September 2022 (25).

## Findings

### emm-type diversity

247 *S pyogenes* isolates were identified from 442 participants between August 2021 and September 2022. 221 isolates were successfully regrown and *emm-*typed. 34 isolates were excluded from population-based analysis to avoid inclusion of multiple isolates coming from the same infectious/carriage episode (see methods), and 187 isolates were included in population-based analyses. 101 isolates were linked with infection (101/187, 54%; 15 pharyngitis, 86 pyoderma) and 86 isolates were linked with carriage (86/187, 46%; 44 pharyngeal carriage, 42 skin carriage) (figure S1). Most isolates were collected in children <18 years (166/187, 88.7%) and in males (124/187, 66.3%). We identified 52 different *emm*-types (57 different *emm*-subtypes) (figure 1b). No novel *emm*-types were found, but 3 new sequences were identified by the CDC as new *emm*-subtypes (28) (table S2). Sixteen *emm*-clusters were identified with most *emm*-types belonging to 4 main *emm*-clusters (65.2%, 122/187), distributed in decreasing order as follow: E3 (20.3%, 38/187), followed by E4 (16.0%, 30/187), D4 (15.5%, 29/187) and E6 (13.4%, 25/187) (figure 1a). A substantial proportion of isolates, 12.8% (24/187), belonged to *emm*-types not classified within any *emm*-cluster (figure 1a, table S2).

**Figure 1.**
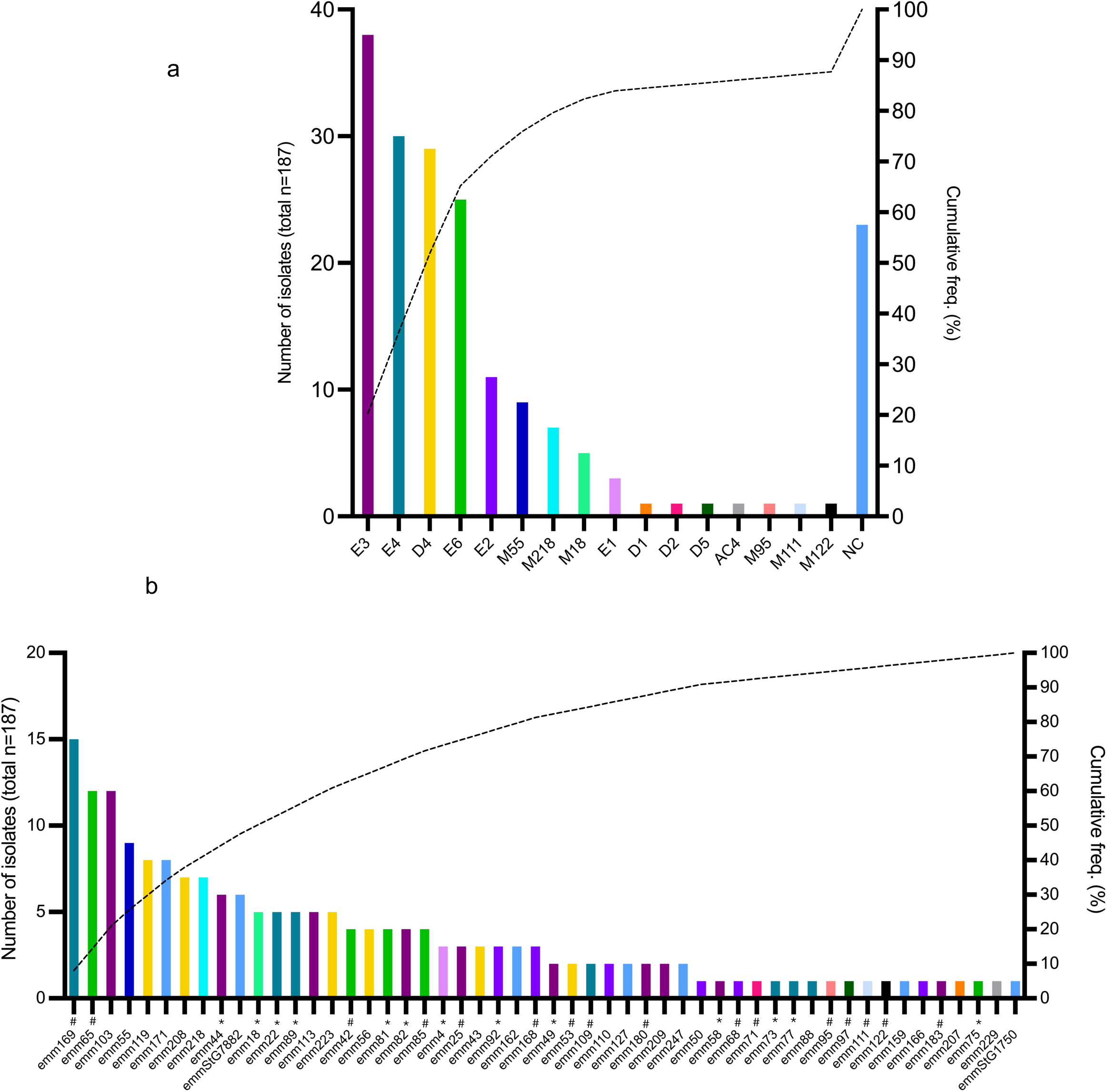
Summary of *emm*-cluster and *emm*-type distribution. **(a)** Summary of 16 different *emm*-clusters distribution. NC stands for “non-classified for *emm*-cluster”, representing the isolates belonging to *emm*-types not classified in any *emm*-cluster. **(b)** Summary of 52 different *emm*-types distribution**. \*** Stands for *emm*-types included in the 30-valent vaccine (21.9%, n=41). **#** Stands for *emm*-types cross opsonised by the 30-valent vaccine (28.9%, n=54). *Emm*-types belonging to the same *emm*-cluster are highlighted with an *emm*-cluster-specific coulour (table S2 for details). Cumulative freq. (%) represents cumulative frequence of isolates in percent.

Simpson’s reciprocal index (SRI) of diversity of *emm*-types over all isolates was 29.3 (95% CI: 24.8-36.0), indicating high strain diversity. Highest diversity was seen in pyoderma (SRI 28.5, 95% CI: 23.4-36.3) and lowest in pharyngitis (SRI 8.3, 95%CI: 5.9-14.4) (table 1). We found no significant differences in *emm*-types or *emm*-cluster regarding tissue tropism or season distribution (figure 2a and b).

**Table 1.** Simpson’s reciprocal index of diversity by isolate type.

| Isolate type | Number of isolates<br>(% total) | Number of different<br>*emm*-types | Simpson’s reciprocal<br>index (95% CI) |
| --- | --- | --- | --- |
| Pharyngeal carriage | 44 (23.5) | 25 | 17.6 (13.0-27.3) |
| Skin carriage | 42 (22.5) | 27 | 18.0 (13.8-26.0) |
| Pharyngitis | 15 (8.0) | 10 | 8.3 (5.9-14.4) |
| Pyoderma | 86 (46.0) | 40 | 28.5 (23.4-36.3) |
| **All isolates** | **187** | **52** | **29.3 (24.8-36.0)** |

**Figure 2.**
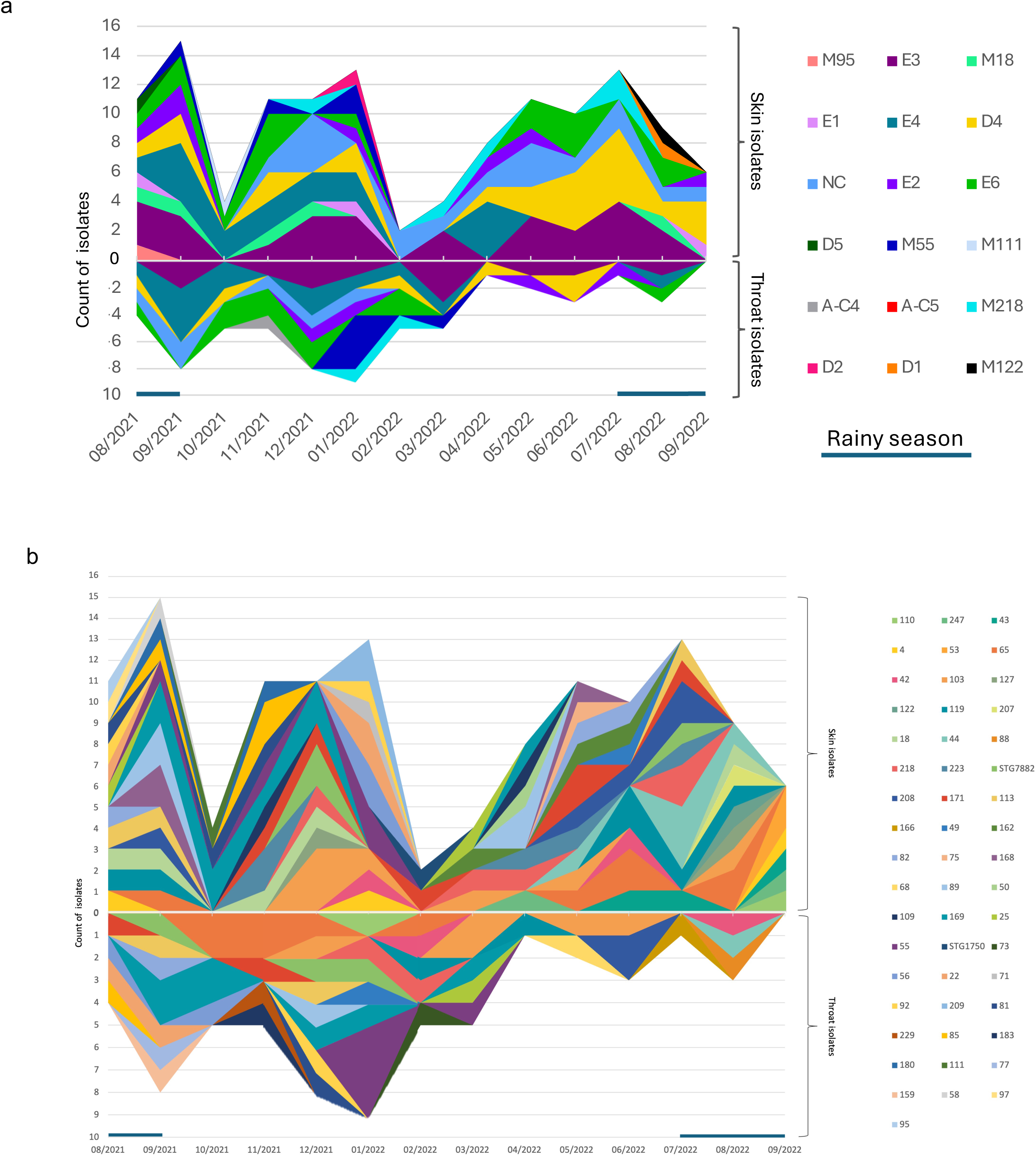
Distribution of *emm*-clusters and *emm*-types over months. Distribution of *emm*-clusters **(a)** and *emm*-types **(b)** over months. Isolates are facetted according to skin isolates (n=128), including pyoderma and skin carriage (top) and throat isolates (n=59), including pharyngitis and throat carriage (bottom). Blue line represents rainy season (July to September). NC stands for “non-classified for *emm*-cluster”. One single isolate collected on 27/07/21 was pooled with august 2021.

### M-protein theoretical vaccine coverage

21.9% (41/187, 95% CI: 16.0-27.8) of our isolates would be directly covered with the 30-valent M vaccine, with an additional 28.9% (54/187, 95% CI: 22.4-35.4) known to be cross opsonised (killing level >50% by functional bactericidal assays) (20, 21) (figure 1b, table S2). Therefore, the potential coverage of the 30-valent vaccine in our cohort is expected to be 50.8% (95/187, 95% CI: 43.6-58.0). 10.7% (20/187, 95% CI 6.3-15.1) of our isolates has been tested and showed no cross opsonisation while 38.5% (72/187, 95%CI: 31.5-45.5) of our isolates have an unknown cross opsonisation (table S2). Regarding the 32-valent vaccine candidate, based on composition vaccine data 18.2% (34/187, 95% CI: 12.6-23.7) of our isolates belong to vaccine sero-types. Theoretical coverage of this candidate could not be estimated due to the absence of clear killing cutoff regarding bactericidal activity (30).

### Temporal comparison of emm-types causing pyoderma

The 2021-2022 pyoderma collection of 86 isolates, comprising 40 *emm*-types, was compared to a 2018 collection of 153 pyoderma isolates, comprising 52 *emm*-types (figure 3a and b) (33). The two collections were sampled in the same neighborhood, 4 years apart. The strains related to pyoderma showed a high diversity in both periods, with an SRI of 35.3 in 2018 (95% IC 30.1-42.7) and 29.1 in 2021-2022 (95% IC 24.2-36.5). More than half of the isolates (135/239, 56.5%) belonged to *emm*-types found in the two periods, while 31.8% (76/239) and 11.7% (28/239) belong to *emm*-types found only in 2018 and 2021-22 respectively (figure 3b). Circulating clusters were broadly stable across both periods with 87.2% of isolates belonging to clusters found in both 2018 and 2021-22 (190/218, excluding the 21 isolates not classified in any cluster) (figure 3a). The 12.8% (28/218) of isolates belonging to clusters found in only one of the two periods do not indicate the emergence of a new emm-phylum (figure S2). The four major *emm*-clusters (E3, E4, D4 and E6), were overrepresented in both periods, accounting for 64.2% of the total isolates (140/218).

**Figure 3.**
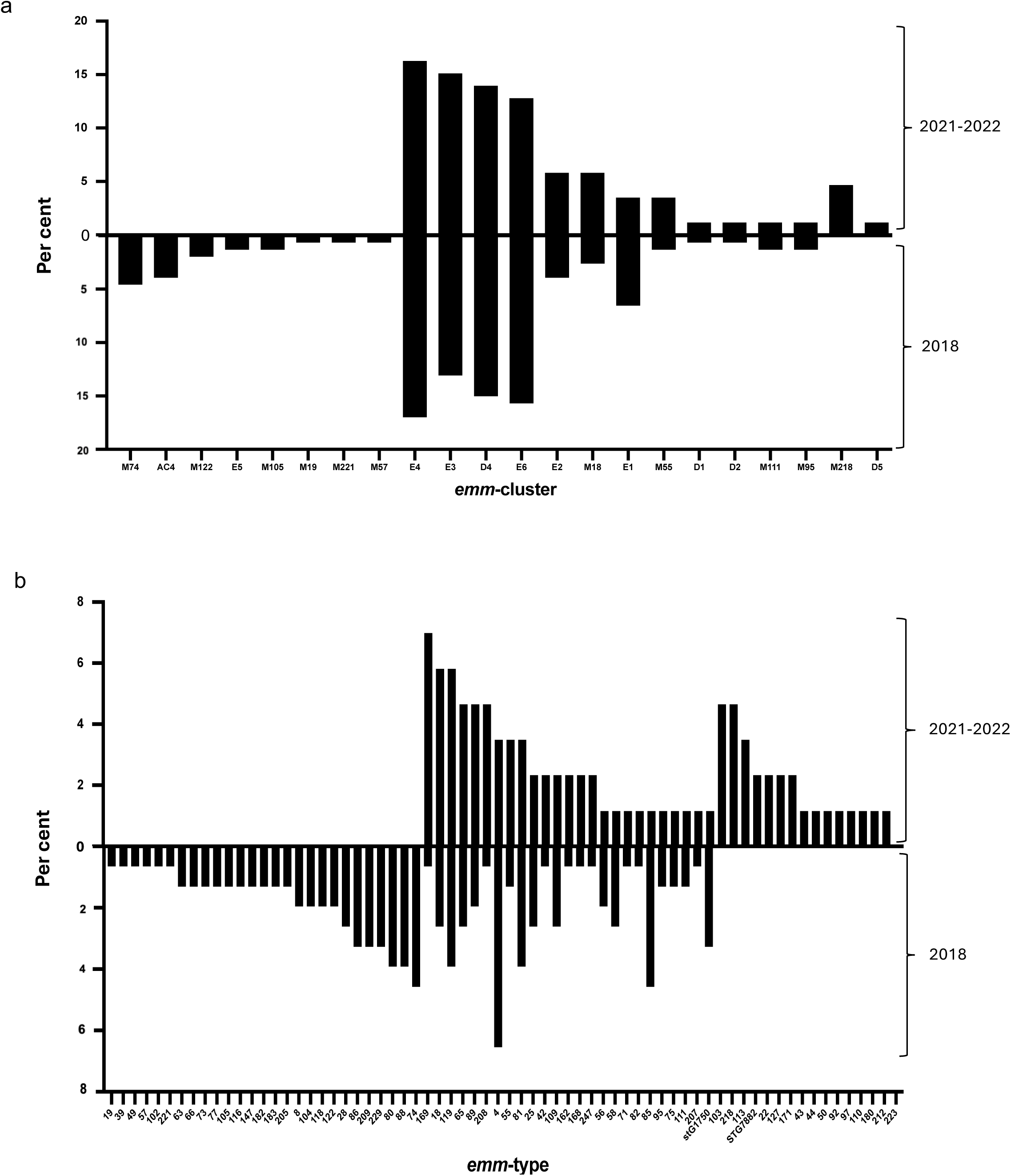
Temporal comparison of *emm*-types and *emm*-clusters associated with pyoderma. Distribution of *emm*-cluster **(a)** and *emm*-types **(b)** associated with pyoderma isolates by collection year. Data presented as percentage of the total number of pyoderma isolates in each collection period (n= 153 for 2018, n=86 for 2021-2022).

### Intra-individual transmission

To assess intra-individual transmission, we focused on multiple isolates collected from the same host within a 31-day interval (figure 4a and b). We selected the 34 participants who had at least two positive isolates collected within a 31-day window (figure 4b). Focusing on intra-individual events due to the same *emm*-type (green arrows, figure 4a), we found frequent transmission between pyoderma and intact skin, and evidence of bidirectional transmission between skin and pharynx in the same host. Pyoderma occurred simultaneously with skin carriage on 8 occasions, and with pharyngeal carriage on 1 occasion. Skin carriage preceded pyoderma on 2 occasions, and pharyngeal carriage on 2 occasions. Pyoderma preceded pharyngeal carriage on one occasion. We did not identify any occasions on which an individual was identified as a pharyngeal carrier and then developed a pharyngitis. Pharyngitis was followed by pharyngeal carriage on two occasions. Persistent skin and pharyngeal carriage with the same *emm*-type were seen on 7 and 11 occasions respectively. The analysis of intra-individual events due to different *emm*-types (red dashed arrows, figure 4b), highlights that different *emm*-types can be carried simultaneously, or a few days apart. This suggests that what appears to be a continuous clinical event based on culture may actually involve different *emm*-types when microbiological analysis is performed. Two individuals presented with concurrent pharyngeal carriage and pyoderma due to different *emm*-types (figure 4a). Empirical treatment of pharyngitis and pyoderma as previously described (26) is likely to have influenced these intra-individual transmission dynamics.

**Figure 4.**
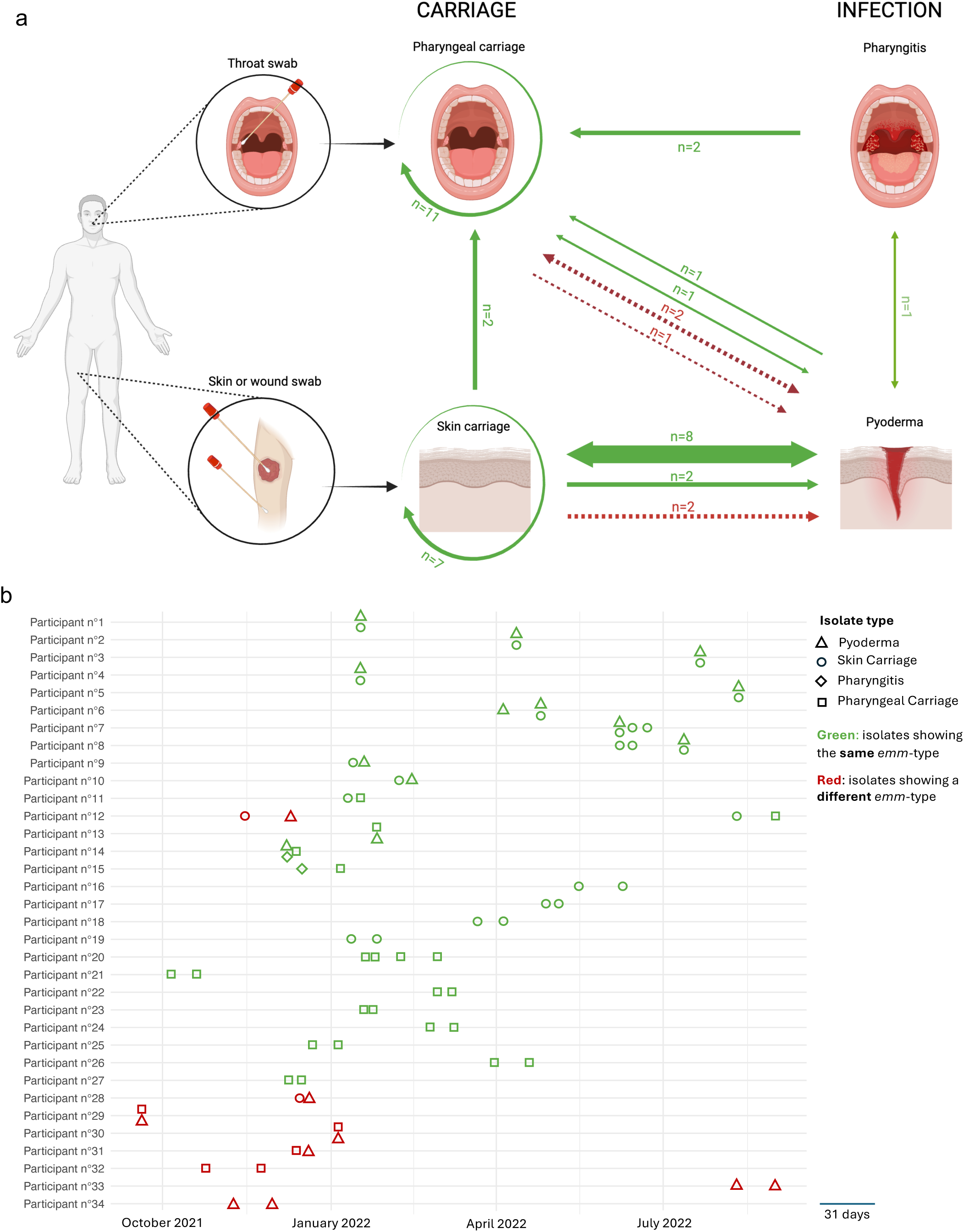
Intra-individual transmission: summary of events due to the same *emm*-type (green) and to a different *emm*-type (red), within a 31-day interval. Schematic representation of intra-individual events happening within a 31-day window. **(a)** The green arrows represent intra-individual events due to the same *emm*-type. Red dashed arrows represent intra-individual events due to different *emm*-types. “n” represents the frequence of each event. The two-way arrows represent events happening simultaneously (on the same day). When not simultaneous, the direction of the arrows indicates which event occurred first. Circular arrows represent persistent skin and pharyngeal carriage. Created in BioRender https://BioRender.com/b75k966 **(b)** Multiple isolates collected from the same participant within a 31-day interval are represented according to their collection date. Isolates collected the same day are stacked. Each point represents an isolate cultured from an individual with pyoderma, skin carriage, pharyngitis or pharyngeal carriage. The points are colored in green if the culture showed the same *emm*-type and red if it showed a different *emm*-type.

## Interpretation

To the best of our knowledge, this is the first molecular epidemiological study to prospectively investigate *S pyogenes* skin carriage in Africa. A recent systematic review of 74 468 isolates originating from 55 countries highlighted the relative lack of epidemiological data coming from Africa, despite being the continent with the highest burden of *S pyogenes* disease and the highest strain diversity (8). It also highlighted the lack of skin isolates in the worldwide literature, representing only 3% of global isolates. Most skin isolates described in the literature are from HIC, with only 3 African studies describing skin infections, representing only 0.2% of global isolates worldwide, and none of these describing skin carriage (table S1) (8). Our study contributes to addressing some of these knowledge gaps by providing robust epidemiological data on skin carriage and infection from The Gambia. Our data highlight the importance of skin infections in this setting, with five times more episodes of pyoderma than pharyngitis. Immune priming by repeated *S pyogenes* skin and throat infections is thought to trigger ARF, and recent studies suggest that, rather than “rheumatogenic” strains driving disease (34), repeated exposure to high strain diversity may lead to a loss of tolerance and autoimmunity in susceptible individuals (12, 35). Pyoderma was associated with the highest strain diversity in our cohort compared to other events, supporting this hypothesis and highlighting the importance of skin infections in the pathogenesis of RHD. To our knowledge, this greater *emm*-type diversity in pyoderma isolates compared to other events is relevant and hasn’t been described before. However, diversity data should be interpreted cautiously, given the small number of pharyngitis isolates compared to other events.

A recent cohort study demonstrated that throat carriage is a reservoir of *S pyogenes* isolates capable of causing pyoderma in remote Aboriginal communities (14). Previous work has demonstrated the importance of skin carriage and pyoderma in inter-individual household transmission in The Gambia (25). Our data suggest frequent transmission between intact skin and pyoderma in the same individual, with evidence of bidirectional transmission between skin and throat in the same host. Our data imply that in this population, the skin could represent the portal of entry into a person and be a major source of transmission. Public health interventions should focus on reducing skin carriage, which appears to be a major reservoir for S pyogenes infections. Consequently, interventions targeting only symptomatic infections could overlook a reservoir of carriers within the population.

Contrary to long-standing beliefs, there is increasing evidence that asymptomatic carriage may be immunologically important (15–17). Immunological studies have so far only considered immune responses to pharyngeal carriage, and responses to skin carriage should also be investigated. Whether carriage poses a significant risk of post-streptococcal sequelae remains to be investigated.

We observed a high diversity of *S pyogenes emm*-types in our cohort, in agreement with previous studies (32, 33, 36). Although three new subtypes were identified, no new *emm*-type was discovered, suggesting that the CDC reference laboratory may have achieved good coverage in terms of global sequence diversity in this area. However, our molecular data highlight the need to include African strains in the *emm*-cluster classification with 12.3% of the isolates that were non-typable with this method (10).

Most isolates (65.2%) belonged to one of the four predominant *emm*-clusters (E3, E4, E6, D4), which are similar to those reported in previous African studies and in low-resource settings worldwide (8, 37). In our temporal comparison, we found little epidemiological change when comparing the 2018 and 2021-22 pyoderma collections. Circulating *emm*-types showed differences, but 87.2% of isolates belonged to *emm*-clusters found in both periods. Any vaccine with broad coverage of the four main clusters circulating in low-resource settings (E3, E4, E6, D4) could therefore be a good strategy, providing protection against most circulating strains in settings with high strain diversity.

Our study has several limitations. First, although the prospective longitudinal design allowed for intensive sampling, we may have missed infection and carriage events in our cohort and prompt treatment may have suppressed transmission and future events. Second, we relied on culture, which is known to be less sensitive than molecular testing (33). Third, although we found a significant number of presumed intra-individual transmission events based on identical *emm*-types, *emm*-typing does not fully distinguish between lineages when compared to whole genome sequencing. Some intra-individual events considered to be *emm*-type related may therefore represent the introduction of separate lineages of the same *emm*-type.

In conclusion, our study highlights the epidemiological relevance of skin infections and skin carriage as a reservoir of *S Pyogenes* in this high RHD burden setting. Strain diversity was high, with the highest diversity seen in pyoderma and the lowest in pharyngitis. However, *emm*-clusters showed lower diversity and appeared to be stable over time, with 4 main clusters gathering 2/3 of the isolates, therefore representing vaccine antigen priorities for broad vaccine coverage. Coordinated efforts to build capacity and surveillance nodes in Africa, with a specific focus on skin infection and asymptomatic carriage, are essential to define *S Pyogenes* transmission chains, inform future preventive measures and support vaccine development.

## Data Availability

All data produced in the present study are available upon reasonable request to the authors.

## Authors’ contribution

SpyCATS study design: TdS, MMarks, APE, AJK. Clinical sample collection: MJ, AB, HC, IC, BS, MM, FF, FM, GdC, APE, AJK. Bacterial identification at MRCG: ES, FC, AJK, GdC. *emm*-typing: GdC, GB, CT, DL, VD, ABO, SB, JH. Data analysis: GdC, LS, PRS, ABO. Writing original Draft: GdC and PRS. Review and Editing: ABO, TdS, APE, AJK, MMarks, CT. All authors revised and approved the final version of the manuscript.

## Informed Consent Statement

The study was approved by the Gambia Government/MRC joint ethics committee and the LSHTM Research Ethics Committee (LEO24005). Written informed consent was provided by adult participants and by parents for participants under 18 years old. Children aged 12-17 provided assent. This study is registered on ClinicalTrials.gov (NCT05117528).

## Acknowledgements

We would like to thank the contribution and strong support of the members of the MRCG Strep A Study Group whose names are not in the main authorship list: Abdul Karim Sesay; Saikou Bah; Beate Kampmann; Annette Erhart; Anna Roca; Isatou Jagne Cox; Peggy-Estelle Tiencheu; Karen Forrest; Sona Jabang; Saffiatou Darboe; Lamin Jaiteh; Aru-Kumba Baldeh; Grant Mackenzie; Martin Antonio. Additional thanks to the MRCG clinical services department led by Karen Forrest for overseeing the clinical care of study participants.

