## Supplementary data for "Household molecular epidemiology of *Streptococcus pyogenes* carriage and infection in The Gambia"

### Supplementary Material

**Table S1.** Systematic literature review of African publications, adapted from Smeesters et al. (1)

| Country | Sampling year(s) | Clinical Manifestations | Nbr isolates included | Nbr pyoderma isolates | Emm-types Simpson's Reciprocal Index (95% CI) | Ref |
|---|---|---|---|---|---|---|
| Ethiopia | 1990 | Pharyngitis, Throat carriage, Skin infection | 214 | 46 | 40.3 (33.7-50) | (2) |
| Ethiopia | 2004-2005 | Throat carriage | 82 | / | NC | (3) |
| Gabon | 2012-2013 | Pharyngitis, Skin infection | 18 | 14 | NC | (4) |
| Kenya | 1998-2011 | Invasive | 328 | / | 53.7 (47.2-62.2) | (5) |
| Mali | 2006-2009 | Pharyngitis | 396 | / | 40 (35.9-45.2) | (6) |
| Morocco | 2017-2018 | Pharyngitis | 13 | / | NC | (7) |
| South Africa | 2008-2001 | Pharyngitis | 150 | / | 14.6 (12.4-17.8) | (8) |
| South Africa | 2017-2018 | Non differentiated | 42 | / | NC | (9) |
| South Africa | 2018-2020 | Non differentiated | 68 | / | NC | (10) |
| South Africa | 2019 | Invasive, Non differentiated | 233 | / | 15.9 (13.2-20.1) | (11) |
| The Gambia | 2004-2018 | Non differentiated | 426 | / | 40.6 (35.7-47.2) | (12) |
| The Gambia | 2018 | Skin infection | 107 | 107 | 33.3 (28.2-40.9) | (13) |
| Tunisia | 2000-2006 | Invasive, Non differentiated | 101 | / | 20.7 (16.6-27.5) | (14) |
| **Total** | | | **2178** | **167** | 65.9 (63.0-69.2) | |

Focus on African publications coming from systematic literature review of global *S pyogenes* strain diversity and disease associations, including 203 articles from 55 countries, covering 74 468 bacterial isolates worldwide. Searches were done in Pubmed, Medline and EmBase from January 1990 until February 2023 (1). The African literature contributes only 3% of global isolates (n=2178), with only 13 studies identified in the worldwide literature, covering 8 African countries. Only 3 African publications include skin isolates (n=167), none of them include skin carriage isolates.

Simpson's Reciprocal Index of diversity is calculated for sampling with more than 100 isolates. NC stands for "not calculated" (sampling with less than 100 isolates). "Non differentiated" refers to isolates for which a clinical type wasn't clearly specified in the referenced article. Nbr: number.

**Supplementary methods: minor methodological differences from previously published work (15):**

As described previously, we can distinguish two subgroups in the main cohort, that underwent more frequent swabbing:

- Clearance time cohort: Participants who acquired new carriage were swabbed from the positive site (oropharyngeal or normal skin) at weekly visits (WVs) until two consecutive negatives swabs to estimate clearance time.
- Intensive sampling cohort: Sixteen randomly selected households (n=160 individuals) underwent weekly intensive visits (IVs) for six weeks during which oropharyngeal and skin swabs were collected, to monitor carriage and infection.

As the clearance time and intensive sampling cohorts underwent more frequent swabbing than the main cohort, carriage acquisitions and infection occurring at those visits were excluded from the incidence analysis in previously published work.

The current work is an epidemiological description of *emm*-types, describing "isolates" rather than "event incidence". Isolates collected at intensive and weekly visits were therefore included in our analysis. However, for population-based analyses, we included a single isolate per *emm*-type per month (31-day interval) and per individual to avoid inclusion of multiple isolates coming from the same infectious/carriage episode, regardless of whether they were collected during WVs or IVs. Multiple isolates of the same *emm*-type collected from the same individual within a 31-day interval were therefore excluded from population-based analyses (*emm*-type diversity, theoretical vaccine coverage and longitudinal comparison). When multiple isolates showing the same *emm*-type were collected from the same individual, those associated with infection were chosen over those linked with carriage. One participant presented with simultaneous pyoderma and pharyngitis due to the same *emm*-type. In this case, both isolates were included, as we did not want to prioritize one infection over the other.

To assess intra-individual transmission, we focused on these multiple isolates belonging to the same *emm*-type collected from the same individual within a 31-day interval (n=34), regardless of whether they were collected during WVs or IVs. The inclusion of WVs and IVs isolates may introduce some bias, as we actively looked for events in participants after carriage or during intensive sampling. Conclusions on the frequency/probability of intra-individual transmission should therefore be drawn with caution.

**Figure S1:** CONSORT flow diagram for isolates included in population-based analysis:

| Main cohort characteristics (n=442*) | |
|---|---|
| **Sex** | |
| Male | 208 (47%) |
| Female | 233 (53%) |
| **Age group** | |
| < 18 years: | 256 (58%) |
| ≥ 18 years: | 185 (42%) |

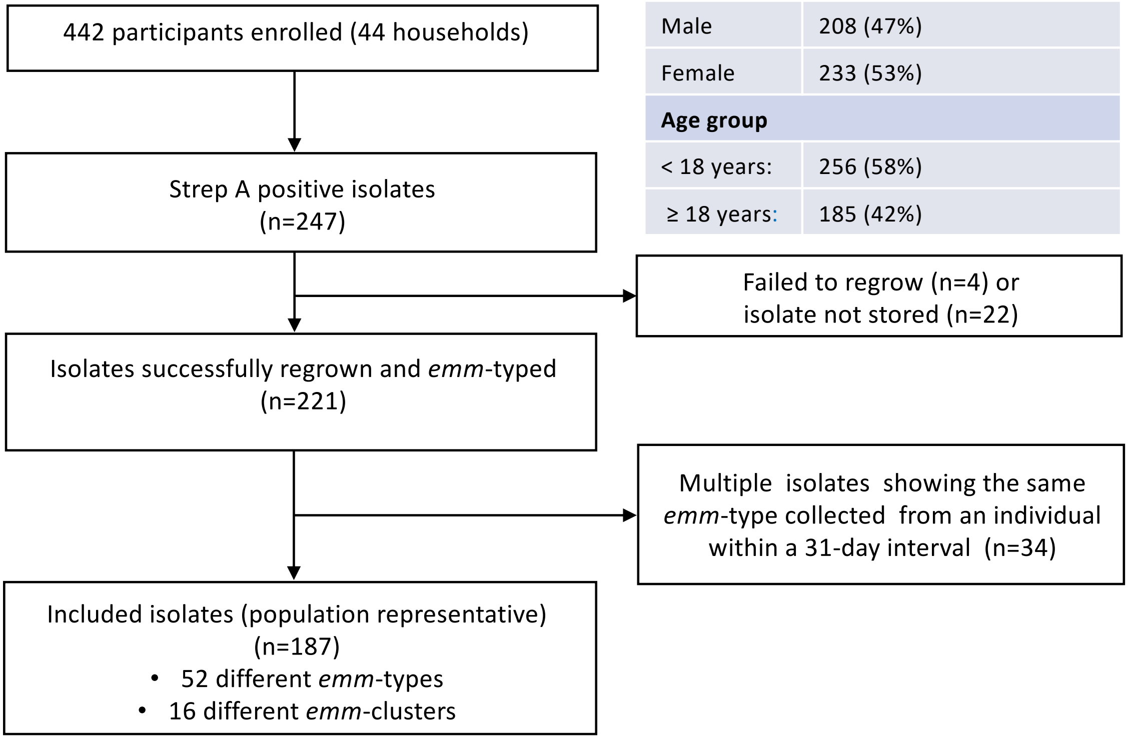


| | Number isolates (n=187) | Total |
|---|---|---|
| **Carriage** | | |
| Skin carriage | 42 (22%) | 86 (46%) |
| Pharyngeal carriage | 44 (24%) | |
| **Infection** | | |
| Pyoderma | 86 (46%) | 101 (54%) |
| Pharyngitis | 15 (8%) | |
| | | 187 (100%) |

*Total cohort was 442 but demographic information was missing for one participant (15)

**Table S2.** Summary of *emm*-types included in population-based analysis and *emm*-cluster distribution:

| *emm*-cluster | Isolates (n) | *emm*-types (n) |
|---|---|---|
| **E3** | 38 | 103.0& (12), 44.0* (6), 113.0& (5), 82.6* (4), 25.1# (3), 49.10* (2), 180.0# (2), 209.3& (2), 58.0* (1), 183.2# (1) |
| **E4** | 30 | 169.1# (15), 22.5* (5), 89.8* (5), 109.1# (2), 73.0* (1), 77.0* (1), 88.11&^ (1) |
| **D4** | 29 | 119.2£ (8), 208.0& (7), 223.0& (5), 56.0& (4), 43.7£ (3), 53.4# (2) |
| **E6** | 25 | 65.7# (10), 81.2* (4), 85.1# (4), 42.0# (2), 42.3# (2), 65.4# (2), 75.3*(1) |
| **E2** | 11 | 92.0* (3), 168.0# (3), 50.0& (1), 68.0# (1), 110.7& (1), 110.10& (1), 166.4& (1) |
| **M55** | 9 | 55.0£ (8), 55.1£ (1) |
| **M218** | 7 | 218.1& (7) |
| **M18** | 5 | 18.21* (5) |
| **E1** | 3 | 4.21* (2), 4.5* (1) |
| **D1** | 1 | 207.0& (1) |
| **D2** | 1 | 71.0# (1) |
| **D5** | 1 | 97.1# (1) |
| **A-C4** | 1 | 229.0& (1) |
| **M95** | 1 | 95.0# (1) |
| **M111** | 1 | 111.4#^ (1) |
| **M122** | 1 | 122.0# (1) |
| **NC** | 23 | 171.1& (8), stG7882.3& (6), 162.1& (3), 127.0& (2), 247.0& (2), 159.3&^ (1), stG1750.1& (1) |
| **Total :** 16 *emm*-clusters | 187 isolates | 52 different *emm*-types (187) |

Table S2. NC stands for “non-classified for *emm*-cluster”, representing the isolates belonging to *emm*-types not classified in any *emm*-cluster. ^ Stands for new subtypes identified in this study (n=3). * Stands for *emm*-types included in the 30-valent vaccine (21.9%, n=41). **#** stands for *emm*-types cross opsonised by 30-valent vaccine (28.9%, n=54). £ Stands for *emm*-types tested and not cross-opsonised by 30-valent vaccine (10.7%, n=20). & Stands for *emm*-types not yet tested for cross opsonisation by 30-valent vaccine (38.5%, n=72).

**Figure S2:** Phylogenetic distribution of *emm*-types and *emm*-clusters associated with pyoderma:

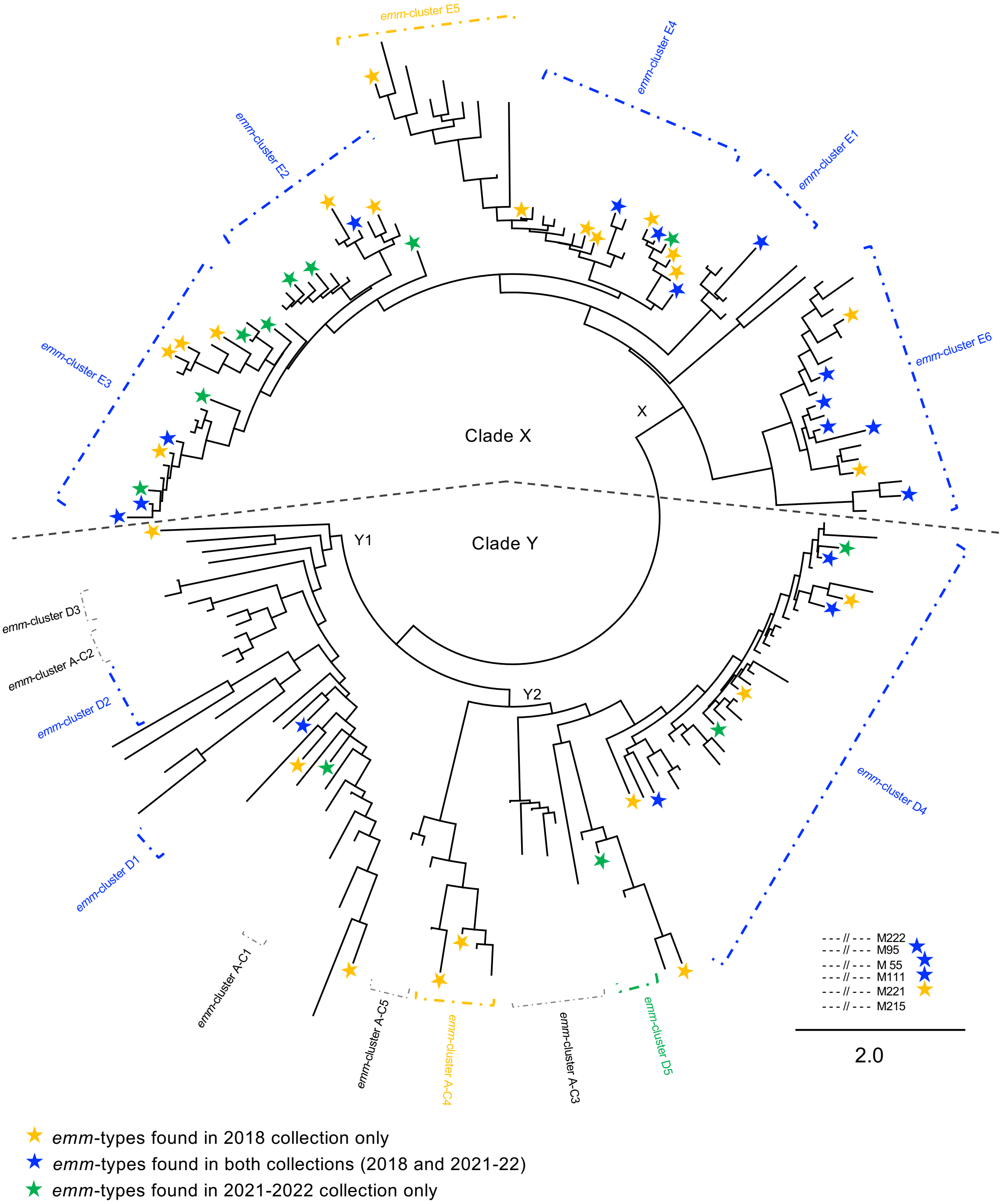


Figure S2. Phylogenetic three based on the M-protein drawn by PhyML, adapted from Sanderson Smith 2014 (16). The three is drawn to scale, branch lengths units are the number of amino acid substitutions per site. The three is supported by an approximate likelihood-ratio test > 80% for the *emm*-clusters as previously described (values are not displayed for clarity reasons). The three is divided in 2 clades: clade X, composed of 6 *emm*-clusters and clade Y, divided in two subclades and composed of 10 emm-clusters. There are 6 outlier emm-types illustrated by dashed lines. Clusters and *emm*-types found only in the 2018 collection are in yellow. Clusters and *emm*-types found in 2018 and 2021-22 are in blue. Clusters and *emm*-types found only in the 2021-22 collection are in green.
